# Environmental and climatic impact on the infection and mortality of SARS-CoV-2 in Peru

**DOI:** 10.1101/2020.09.16.20196170

**Authors:** Victor J. Samillan, Diana Flores, Eduardo Rojas, Brian R. Zutta

## Abstract

The role of the environment and climate in the transmission and case-fatality rates of SARS-CoV-2 is still being investigated. Elevation and air quality are believed to be significant factors in the current development of the pandemic, but the influence of additional environmental factors remain unclear.

In this study, we explored the relationship between the cumulative number of infections and mortality cases with climate (temperature, precipitation, solar radiation, water vapor pressure, wind), environmental data (elevation, NDVI, PM_2.5_ and NO_2_ concentration), and population density in Peru. Using the data from confirmed cases of infection from 1287 districts and confirmed cases of mortality in 479 districts, we used Spearman’s correlations to assess the correlation between environmental and climatic factors with cumulative infection cases, cumulative mortality and case-fatality rate. We also explored district cases by the ecozones of coast, sierra, high montane forest and lowland rainforest.

Multiple linear regression models indicate elevation, mean solar radiation, air quality, population density and green cover are influential factors in the distribution of infection and mortality of SARS-CoV-2 in Peru. Case-fatality rate was weakly associated with elevation. Our results also strongly suggest that exposure to poor air quality is a significant factor in the mortality of individuals with SARS-CoV-2 below the age of 30.

We conclude that environmental and climatic factors do play a significant role in the transmission and case-fatality rates in Peru, however further study is required to see if these relationships are maintained over time.

## Introduction

As SARS-CoV-2 has spread around the world in a few months, several groups of investigators have started to look into the different factors that could be related to the distribution of the infection and the severity of the disease. The incidence of infection in different countries and cities suggests that several factors influence the rate of infection, which are not only related to the virus and the immune system. In the case of Peru, the cities that have the most cases are generally located near the coast and have poor air quality throughout the year (https://www.datosabiertos.gob.pe/). In addition, population density seems to be a key factor in the spread of the virus, where overcrowded cities increase the probability of contact with infected individuals, increasing the number of infection and mortality cases in a short amount of time. Therefore, population density should be taken into account when analyzing different cofactors SARS-CoV-2 distribution.

Although more studies are necessary, the rate of infection and the severity of the diseases seems different for people living in cities at high altitudes, where not only hipoxia is a major factor, but other factors such as air quality, solar radiation, and population density, could play a role in SARS-CoV-2 person-to-person transmission. Arias-Reyes et al. (2020) suggested that there exists less rate of infection at high altitude possibly due to a lower level of expression of ACE2 compared to sea level. This observation is important, particularly since SARS-CoV-2 uses ACE2 as a point of infection for the cells (Ren et al., 2020). Yao et al. (2020) explored the rate of SARS-CoV-2 infection across China, and concluded that there was no correlation between temperature, UV and the rate of infection. However, they recommended future studies using more complex models and environmental factors. Finch et al., (2016) explored the role of pollution exposure in the development of different diseases, where heart and respiratory diseases indicated a detrimental role of pollution on the endothelial integrity, and the changes in the levels of Endothelin I could be observed in people that were exposed to air pollution.(Briet et al. 2007)

Environmentals factors related to mechanics of different virus spread have been studied by several groups, before the pandemic. Pica et al. (2012) studied the environmental factors related to the spread of seasonal influenza and concluded that some meteorological factors play a central role in the person-to-person transmission of the virus, besides the sociodemographic factors. It is well known that UV radiation can decontaminate surfaces and air, and UV incidence increases at higher elevations and cities will have different exposures depending on their location. Kowalsky et al. (2009) demonstrated that the UV light can destroy a bacteries, many viruses and fungi, but additional studies are necessary to understand the impact of UV on SARS-CoV-2 in the natural environment.

Understanding all the factors that are related to the spread of SARS-CoV-2 will be an important part of public policy, particularly towards the implementation of focalized quarantines in cities and districts where the cases of infection are high. Therefore, this study aims to explore the different cofactors that could increase or decrease the possibility of person-to-person transmission of SARS-CoV-2. The main objectives of this study was to explore the relationship between SARS-CoV-2 infection and mortality cases, case-fatality rates with a set of climate (temperature, precipitation, solar radiation, water vapor pressure, and wind), environmental data (elevation, NDVI, PM_2.5_ and NO_2_ concentration), and population density in Peru. We also divide the distribution of infection, mortality and case-fatality rates by four ecozones (coast, sierra, high montane forest and lowland rainforest) and subset mortality cases by age groups and gender.

## Methods

We obtained SARS CoV 2 confirmed cases by district, of the 1873 districts in Peru, from official reports provided by the Peruvian Ministry of Health (MINSA), through an official government open data portal (https://www.datosabiertos.gob.pe/). We used the cumulative number of confirmed cases of infection from 1287 districts and confirmed cases of mortality in 479 districts, where at least 1 confirmed case was registered as of June 27, 2020 (Figure 1). The first case of SARS CoV 2 was registered on March 5, 2020. Under this criteria, the number of positive SARS CoV 2 cases analyzed were 263,743 and 7,877 deaths by June 27, 2020.

**Figure 1.**
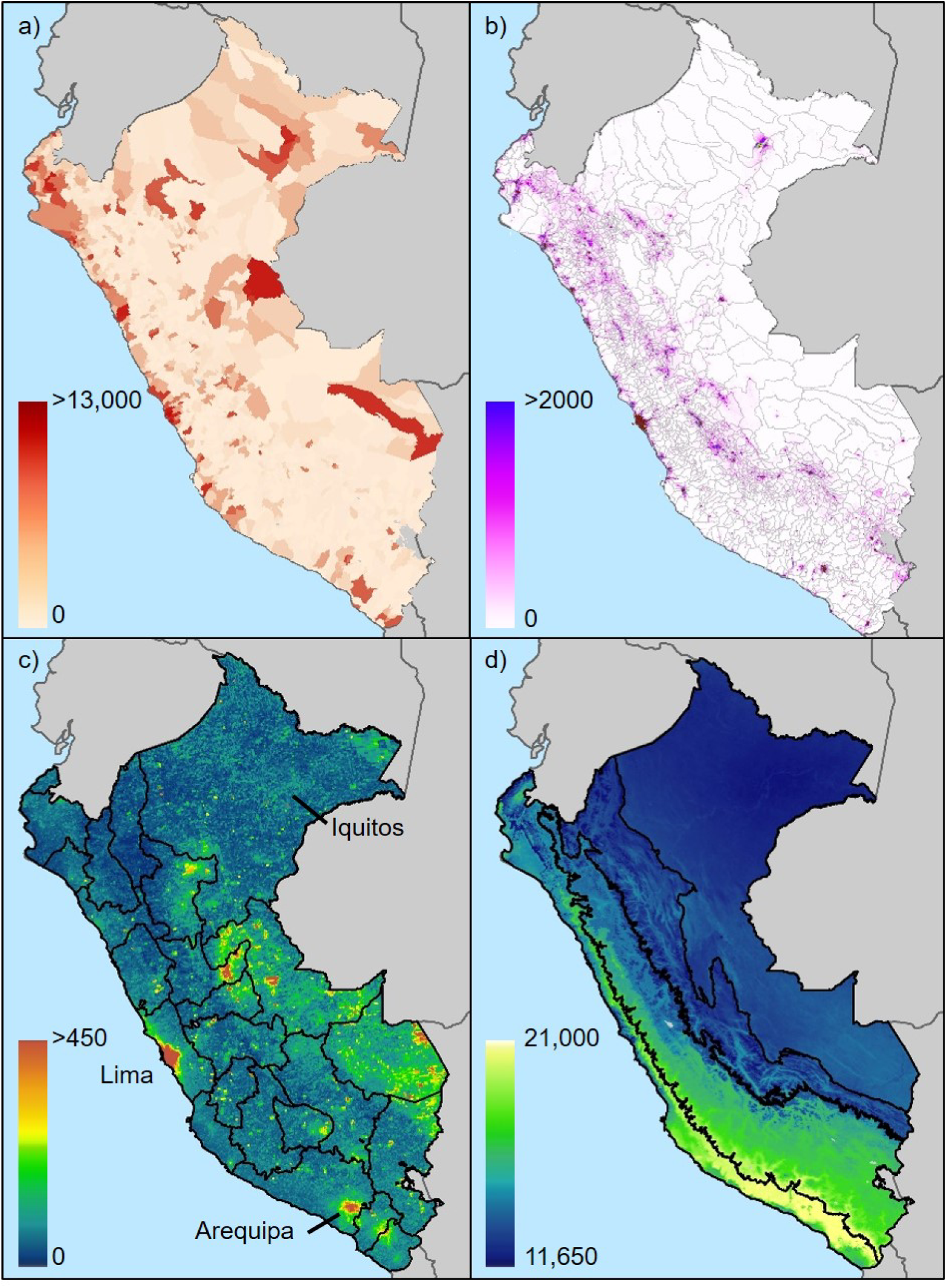
a) Cumulative number of confirmed infected cases per district, b) modeled population density (people km^−2^) for the year 2020, c) maximum tropospheric NO_2_ density (µmol m^−2^) for 2019, and d) mean solar radiation (kJ m^−2^ day^−1^). Outlines in a) and b) are district boundaries, while outlines in c) indicate departmental boundaries and d) (left to right) the coast, sierra, high montane forest and lowland rainforest ecozones.

Available data of infection included date of confirmation, while data on mortality included age, sex, and date registered. The first confirmed case of SARS CoV 2 in Peru was on March 6, 2020. Infection and mortality values were log-transformed to meet statistical assumptions in relation to residuals and added 1 to avoid taking the logarithm of 0 (Liu et al, 2020; Xie and Zhu, 2020; Zhu et al., 2020).

We also assessed case-fatality rates, which implies the severity of the condition by estimating the proportion of cases that die of a given condition (Segovia-Juarez et al., 2020). The case-fatality rate was estimated by dividing the number of cause-specific deaths among the incidence cases by the total number of incident cases*100 (CDC, 2019)

We labeled districts based on four ecozones, including coast, sierra, high montane forest and lowland rainforest, which was originally developed based on elevation and biogeography and used in national and international reporting, and have a significant effect on agricultural activity, population density, population connectivity and other socioeconomic activities (MINAM, 2016).

### Climate and environmental data

A set of climate metrics were obtained from WorldClim version 2.1 (Fick & Hijmans, 2017), which presents a historical baseline from the years 1979 to 2000 and includes monthly temperature, rainfall, wind speed, and solar radiation. Satellite-based environmental data was obtained from a variety of satellite sensors that detect environmental variables, including elevation, vegetation cover, and air quality. Vegetation cover was estimated through NDVI (Normalized Difference Vegetation Index), a commonly used spectral index used to quantify green cover, including agricultural extent, forest cover, and green cover in urban settings. We infer air quality through the metrics of two health-relevant air pollutants, which include particulate matter at 2.5 μm (PM_2.5_) and nitrogen dioxide (NO_2_). Population density was obtained from the WorldPop model of population density (people km^−2^) for Peru adjusted to match official United Nations population estimates for the year 2020 (www.worldpop.org). Further details on climate data and satellite-based environmental data are provided in the online supplementary material.

### Data analysis

We extracted the zonal statistics by district (i.e. average value per district polygon) of each of the climate, remotely sensed and population density layers using ArcPro (verison 2.2). We used SPSS 25.0 (IBM, USA) for all statistical analysis of the extracted values. We used the one-sample Kolmogorov-Smirnov parametric test to evaluate the distribution of residuals obtained for each district from each of the original 36 data layers used. Covariance within the subsets of climate and environmental variables were estimated with Spearman’s Rho correlation, where correlation of the order of 0.9 or larger were determined to have a high covariance. As a result, the final reduced data set used in this study were 21 data layers, including 14 climatic layers, 6 environmental layers and the population density model (Table 1). The climatic layers included 4 temperature, 4 rainfall, 2 solar radiation, 2 wind speed and 2 water vapor metrics. All 6 environmental layers and the population density model had a relatively low level of covariance and were kept for further analysis. The climate and environmental data were also normalized (following Zhu et al., 2020), due to the different units used in each variable, through the following method:

**Table 1.** Description of the variables selected for statistical modeling.

| Data | Source | Temporal resolution | Spatial resolution |
| --- | --- | --- | --- |
| Climatic variables |  |  |  |
| Annual mean temperature | WorldClim | 1970–2000 | 1 km |
| Mean diurnal temperature range | WorldClim | 1970–2000 | 1 km |
| Temperature seasonality | WorldClim | 1970–2000 | 1 km |
| Temperature annual range | WorldClim | 1970–2000 | 1 km |
| Annual precipitation | WorldClim | 1970–2000 | 1 km |
| Precipitation of Wettest Month | WorldClim | 1970–2000 | 1 km |
| Precipitation seasonality | WorldClim | 1970–2000 | 1 km |
| Precipitation of warmest quarter | WorldClim | 1970–2000 | 1 km |
| Solar radiation (mean) | WorldClim | 1970–2000 | 1 km |
| Solar radiation (standard deviation) | WorldClim | 1970–2000 | 1 km |
| Water vapor pressure (mean) | WorldClim | 1970–2000 | 1 km |
| Water vapor pressure (standard deviation) | WorldClim | 1970–2000 | 1 km |
| Wind speed (mean) | WorldClim | 1970–2000 | 1 km |
| Wind speed (standard deviation) | WorldClim | 1970–2000 | 1 km |
| Environmental variables |  |  |  |
| Elevation | SRTM | 2000 | 90 m |
| NDVI (mean) | MODIS | 2019 | 500 m |
| PM 2.5 (mean) | MERRA-2 | 2019 | 62 km |
| Tropospheric NO<sub>2</sub> (mean) | Sentinel-5P | 2019 | 500 m |
| Tropospheric NO<sub>2</sub> (maximum) | Sentinel-5P | 2019 | 500 m |
| Population density | WorldPop | 2020 | 1 km |

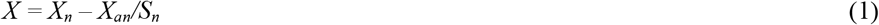

Where *X* is the normalized data, *X*_*n*_ is the raw data for the variable, *X*_*an*_ is the mean value of the raw data, and *S*_n_ is the standard deviation of *X*_*n*_.

We used stepwise linear regression to explore the association of climatic and environmental variables with the log-transformed data of cumulative number of confirmed cases of infection and confirmed cases of mortality, across Peru and within ecozones. The number of cases were used as dependent variables, and the environmental and climatic were selected as independent variables. The resulting statistically significant predictive models use one or multiple variables that best explain the dependent variable. The adjusted R^2^ from model fitting indicates the percentage of all factors that explain the distribution of cases and the magnitude of the standardized β reflects the influence of the corresponding variable in the predictive model. Collinearity diagnostics, resulting from the regression, were also used to identify model variables that were highly correlated. Significant differences between means were evaluated using one-way ANOVA followed by a Tukey test.

## Results

The district of San Juan de Lurigancho, of the department of Lima and in the coastal ecozone, had the most number of confirmed infected cases and case fatalities of SARS-CoV-2, with 13,724 total infections and 394 fatalities, up until the date of the data obtained for this study. The average number of confirmed cases of infection and fatalities was 205 (SD ± 938) and 16.4 (SD ± 42.2) per district, respectively. Across ecozones, the number of registered male fatalities was 5,601 (71.1%) and females was 2,276 (28.9%). The average age of total male fatalities was 64.2 (SD ± 15.6) and females was 66.3 (SD ± 15.9). There was also a significant difference between ecozones for mean age of fatalities (*F* = 5.64, *p*<0.001) and female only cases (*F* = 6.03, *p*<0.001), but not in male only cases (*F* = 1.46, *p*=0.224). We found no significant difference between the mean case-fatality rates by ecozones (*F* = 0.34, *p*=0.80; Table 2).

**Table 2.** Difference between mean age ± SE of fatalities in districts within different ecozones. Min-max represents the range of age and n is the number of cases.

|  | Ecozone |  |  |  |  |
| --- | --- | --- | --- | --- | --- |
| Cases | Coast | Sierra | High montane forest | Lowland rainforest | 1-way ANOVA *p*-value |
| Cumulative fatalities | 65.1 ± 0.2 | 66.5 ± 0.7 | 62.3 ± 1.3 | 62.8 ± 0.8 | < 0.001 |
| min-max | 0–101 | 16–97 | 1–99 | 2–95 |  |
| n | 4737 | 423 | 192 | 390 |  |
| Male fatalities | 64.4 ± 0.3 | 64.7 ± 0.9 | 62.5 ± 1.4 | 62.8 ± 1.0 | 0.224 |
| min-max | 0–101 | 16–97 | 1–90 | 6–95 |  |
| n | 3091 | 281 | 138 | 274 |  |
| Female fatalities | 66.4 ± 0.4 | 70.0 ± 1.2 | 61.6 ± 2.7 | 62.8 ± 1.6 | < 0.001 |
| min-max | 0–100 | 25–95 | 1–99 | 2–92 |  |
| n | 1646 | 142 | 54 | 116 |  |

Correlation analysis indicated a positive correlation of total infections with PM_2.5_ (□ = 0.449) and a negative correlation with elevation (□ = –0.476), where the number of infections correlates with increased air pollution, while there are less number of infections with elevation Figure 2). Similarly, correlation analysis indicated a high positive correlation coefficient between mean tropospheric NO_2_ (□ = 0.497, p<0.001), PM_2.5_ (□ = 0.506; p<0.001) and registered mortality across sexes and in the male population with □ = 0.504 and □ = 0.486, respectively.

**Figure 2.**
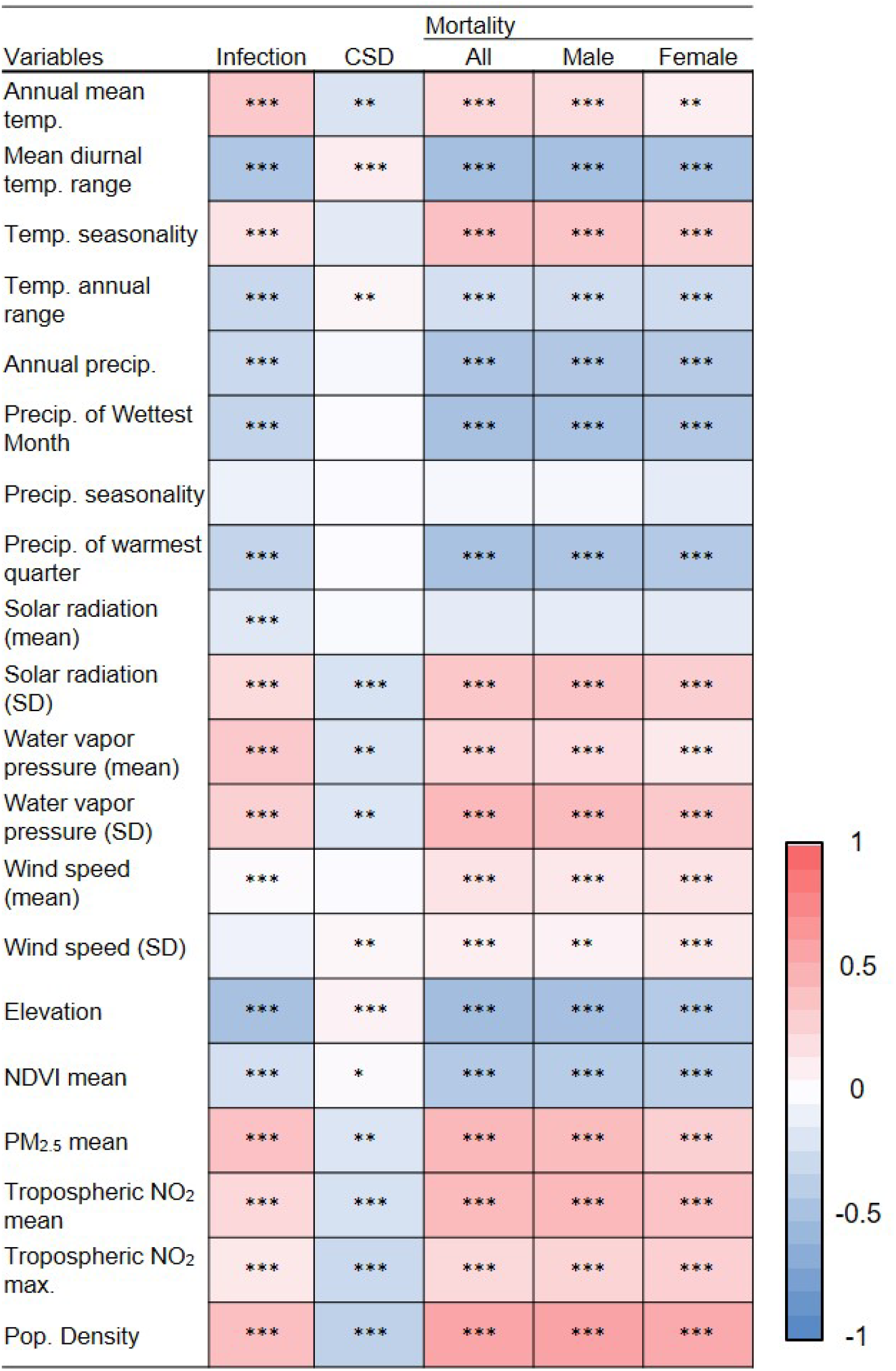
Spearman’s correlation between climatic and environmental factors and the cumulative cases of infection, case-fatality rate (CFR) and mortality. The color gradient indicates correlation coefficients, where darker red indicates a correlation approaching 1 and a darker blue indicates a correlation approaching –1. Statistical significance were * p<0.05, ** p <0.01, and ***p<0.001.

In stepwise linear regression model fitting, the three most important parameters that explain the distribution of cumulative number of infections across the country (R^2^ = 0.46; p-value = 0.038) were NDVI (β = –0.392), elevation (β = –0.311), and mean solar radiation (β = –0.244)(Table 3). The negative correlation indicates a decrease in the cumulative number of infections, with an increase in surrounding green cover (NDVI). In addition, the cumulative number of infections decreases with increasing elevation and increasing mean solar radiation. Within each ecozone, parameters importance change indicating the importance of population density (β = 0.474) in the coast, NDVI (β = –0.359) in the sierra, maximum NO_2_ (β = 0.352) in high montane forest, and elevation (β = –0.441) in lowland rainforest.

**Table 3.** Predictive models of the relationship of cumulative case infection and environmental and climatic variables.

| Dependent | Predictors | Standardized $\beta$ | $R^2$ | Adj. $R^2$ | $p$-value |
| --- | --- | --- | --- | --- | --- |
| Cumulative inf. | NDVI | -0.392 | 0.46 | 0.46 | 0.04 |
|  | Elevation | -0.311 |  |  |  |
|  | Radiation mean | -0.244 |  |  |  |
|  | NO<sub>2</sub> max | 0.171 |  |  |  |
|  | NO<sub>2</sub> mean | 0.057 |  |  |  |
|  | PM<sub>2.5</sub> | 0.129 |  |  |  |
|  | Pop. density | 0.150 |  |  |  |
|  | Wind mean | -0.081 |  |  |  |
|  | Radiation (SD) | -0.077 |  |  |  |
| Coast | Pop. density | 0.474 | 0.35<br>0 | 0.34 | 0.03 |
|  | Elevation | -0.199 |  |  |  |
|  | PM2.5 | 0.163 |  |  |  |
| Sierra | NDVI | -0.359 | 0.50 | 0.50 | 0.02 |
|  | Radiation mean | -0.272 |  |  |  |
|  | Elevation | -0.244 |  |  |  |
|  | NO<sub>2</sub> mean | 0.191 |  |  |  |
|  | PM<sub>2.5</sub> | 0.171 |  |  |  |
|  | Pop. density | 0.131 |  |  |  |
|  | Wind SD | -0.077 |  |  |  |
| High montane rainforest | NO<sub>2</sub> max | 0.325 | 0.40 | 0.39 | <0.001 |
|  | PM<sub>2.5</sub> | 0.266 |  |  |  |
|  | Elevation | -0.253 |  |  |  |
| Lowland rainforest | Elevation | -0.441 | 0.37 | 0.35 | 0.001 |
|  | Pop. density | 0.327 |  |  |  |

Model fitting for cumulative mortality cases across the Peru indicated the negative correlation of NDVI (β = –0.299), elevation (β = –0.245), and mean solar radiation (β = –0.230), and the positive correlation with mean NO_2_ (β = 0.237) and population density (β = 0.215) (Table 4). Population density was the most influential factor for mortality in the coastal ecozone, lowland rainforest, and male and female mortality across the country. It is worth noting that adjusted R^2^ indicated that the models included at least 28% of the factors that affect the difference in mortality.

**Table 4.**
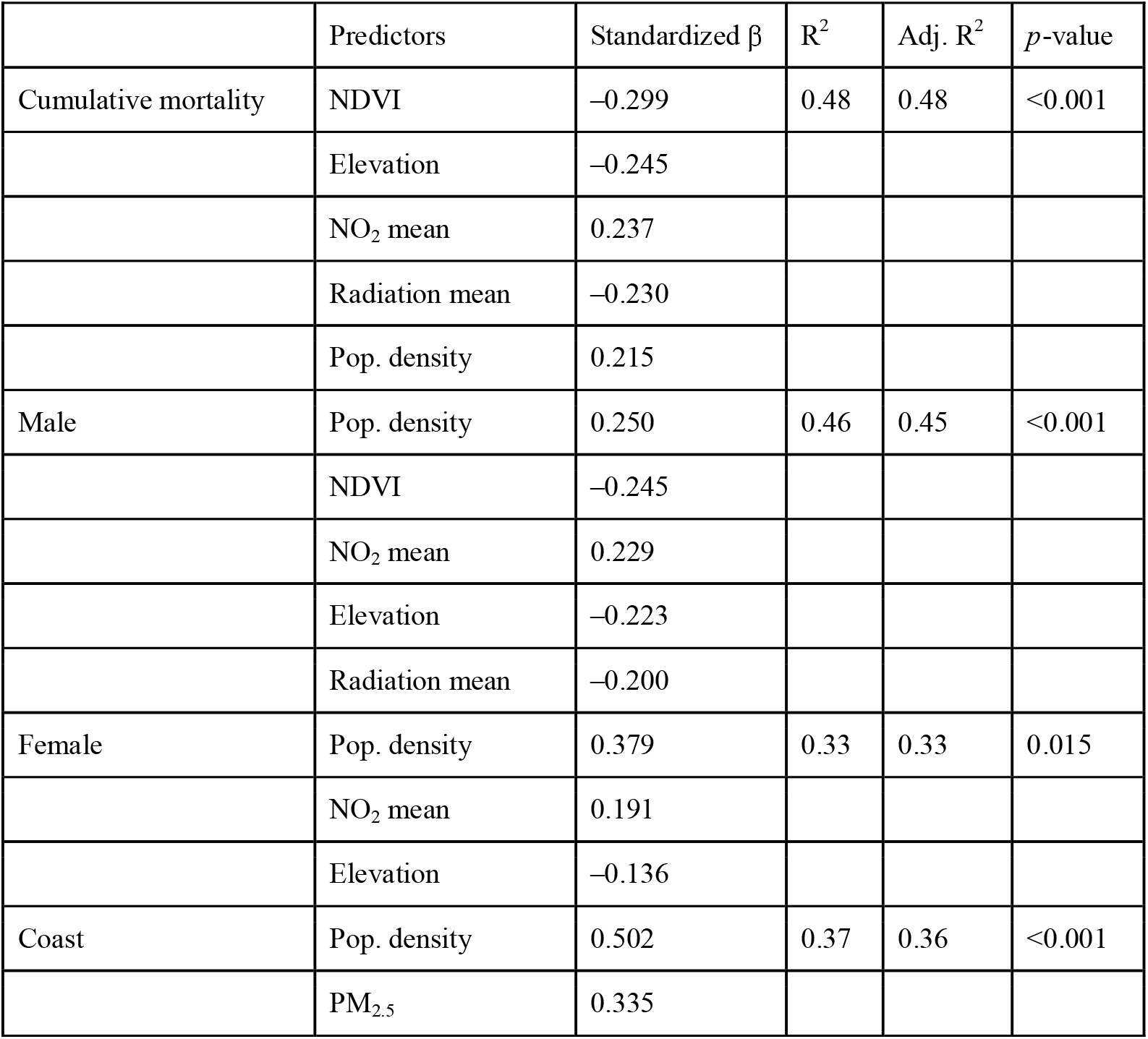
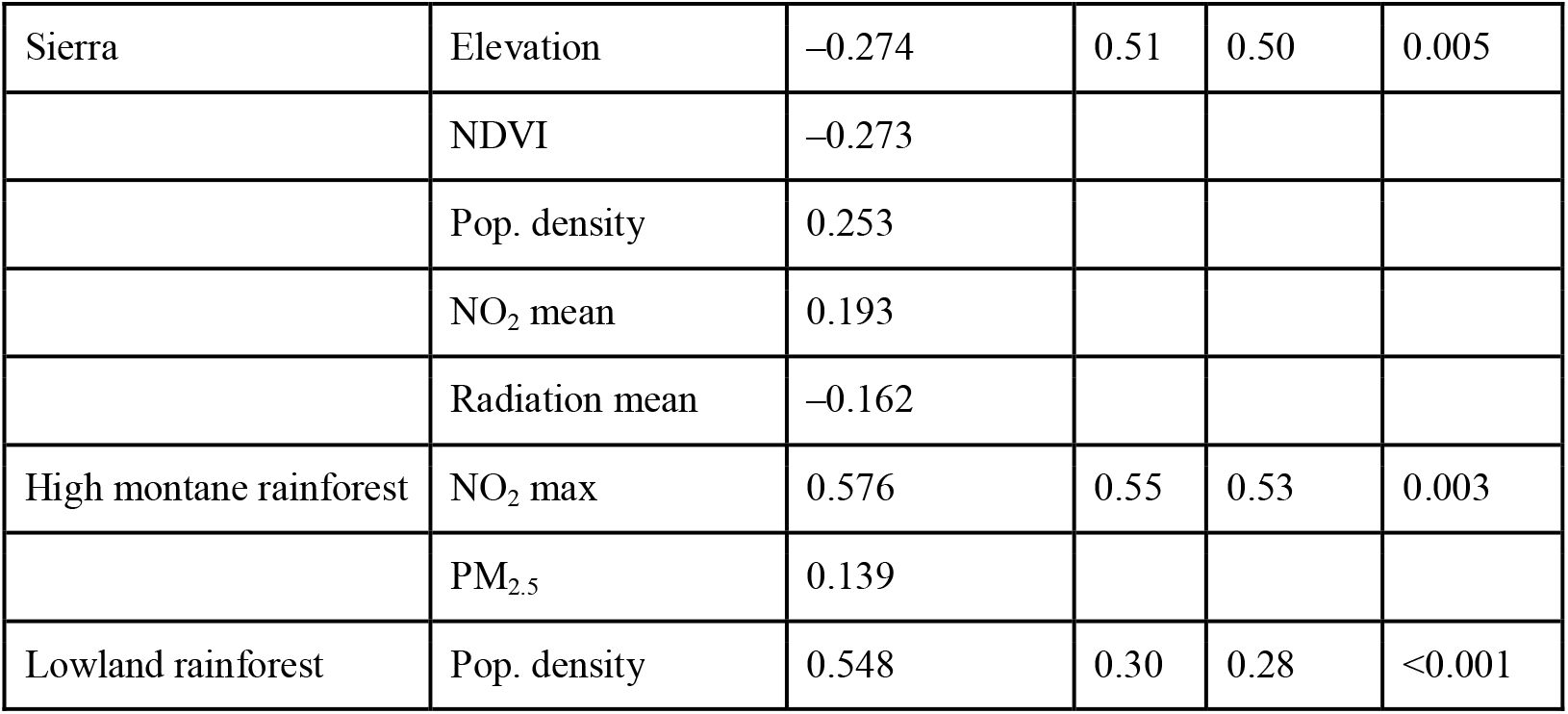
Predictive models of the relationship of mortality across and within ecozones and environmental and climatic variables.

Regarding age groups, mean NO_2_ was the most influential factor of difference of mortality for individuals with an age range of 0 to 17 (β = 0.654) and 18 to 29 (β = 0.422) (Table 5). Population density was the most influential factor for the remaining age ranges of 30 to 49 (β = 0.306), 50 to 70 (β = 0.483), and above 80 (β = 0.309). However, it is worth noting that mean NO_2_ was indicated as an influential factor for mortality for the age ranges of 30 to 49 (β = 0.236) and above 80 (β = 0.258).

**Table 5.**
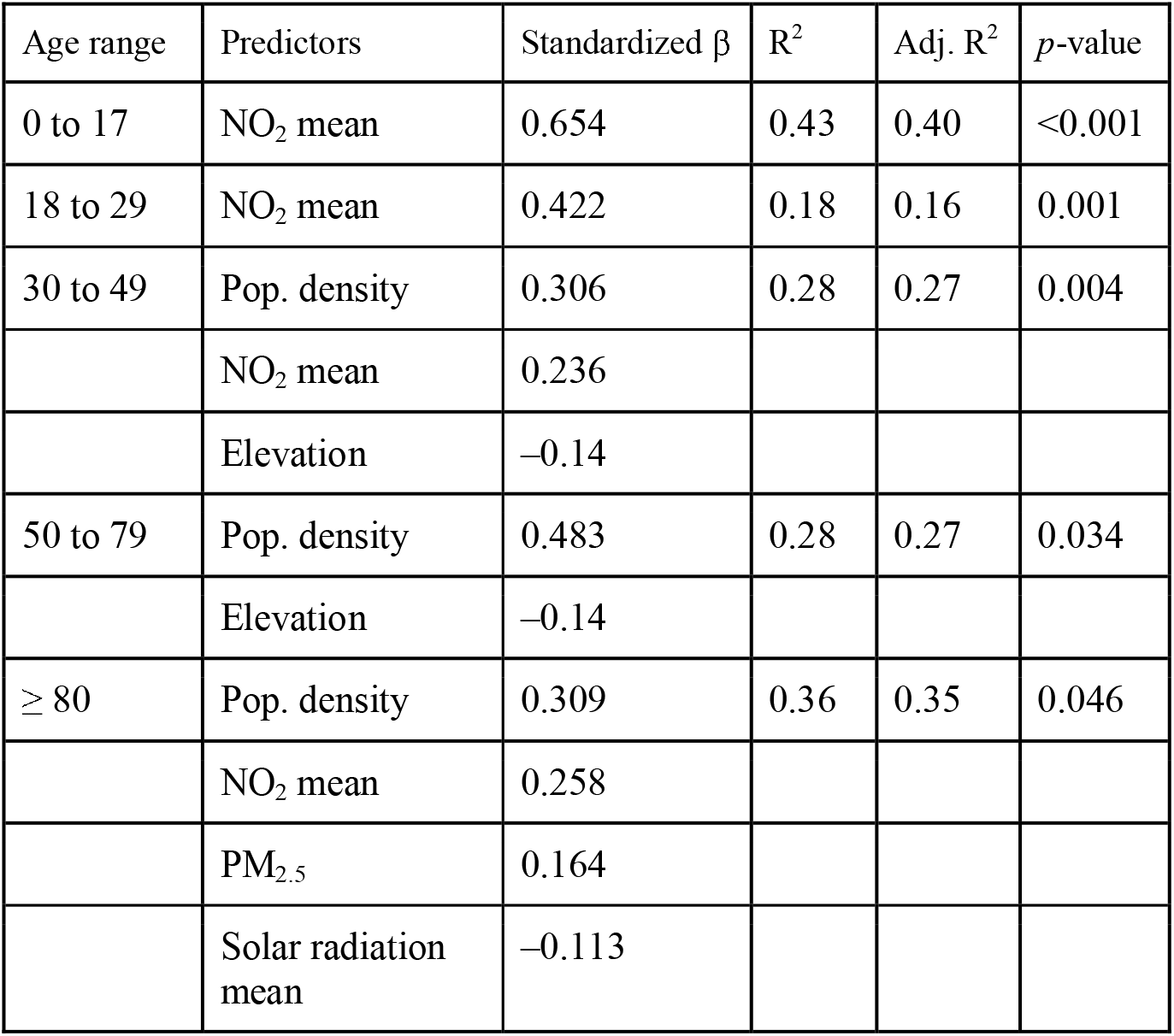
Predictive models of the relationship of age of mortality and environmental and climatic variables.

A positive correlation from model fitting was found between case fatality rates and elevation (β = 0.318), although a relatively low explanatory factor (R^2^ = 0.12; p-value = 0.015) compared to most predictive models performed in this study (Table 6). Indeed, case fatality rate models had the lowest R^2^, although statistically significant (p < 0.05), compared to models for infection and mortality. Elevation was the most influential factor for the coast (β = 0.281), sierra (β = 0.303), and high montane forest (β = 0.466). Mean diurnal range was the main factor for case fatality rates in high montane forests (β = 0.466) and the only climate variable to appear as a factor for model fitting in this study.

**Table 6.**
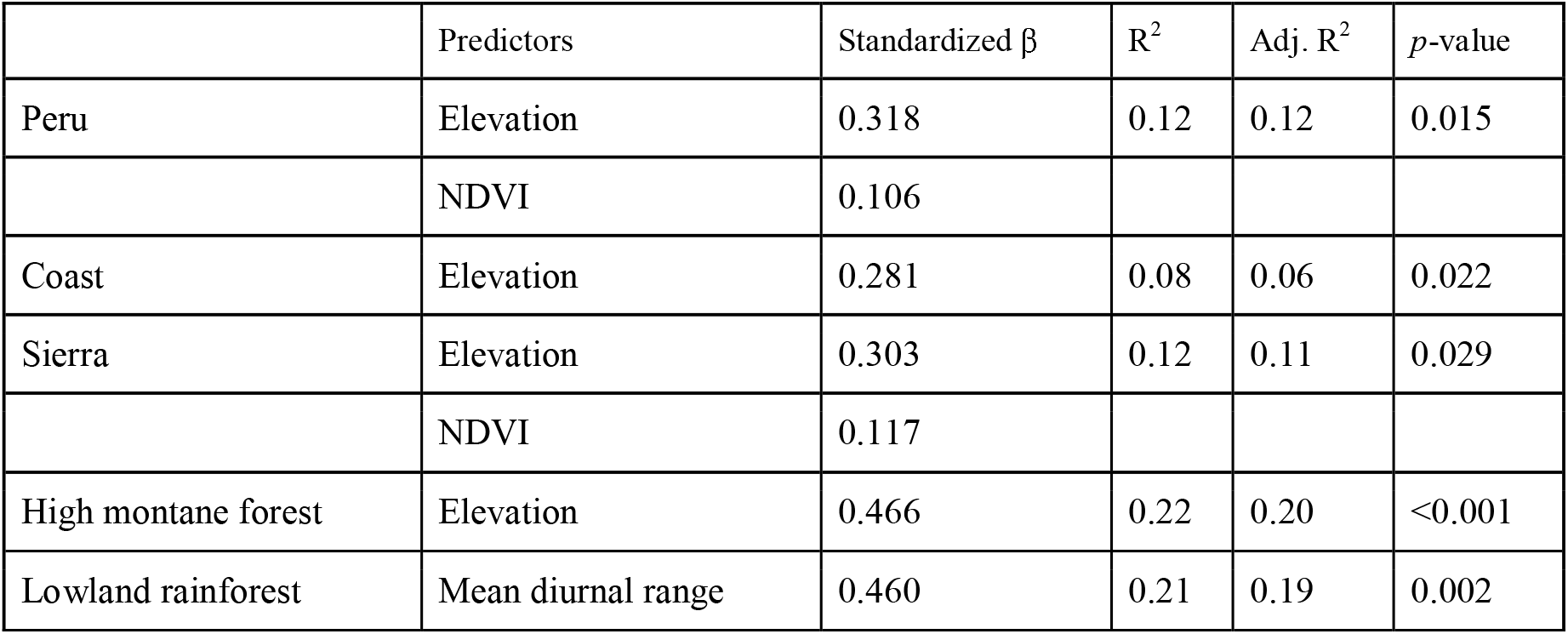
Predictive models of the relationship of case fatality rates and environmental and climatic variables.

| | Predictors | Standardized $\beta$ | $R^2$ | Adj. $R^2$ | $p$-value |
| --- | --- | --- | --- | --- | --- |
| Peru | Elevation | 0.318 | 0.12 | 0.12 | 0.015 |
|  | NDVI | 0.106 |  |  |  |
| Coast | Elevation | 0.281 | 0.08 | 0.06 | 0.022 |
| Sierra | Elevation | 0.303 | 0.12 | 0.11 | 0.029 |
|  | NDVI | 0.117 |  |  |  |
| High montane forest | Elevation | 0.466 | 0.22 | 0.20 | <0.001 |
| Lowland rainforest | Mean diurnal range | 0.460 | 0.21 | 0.19 | 0.002 |

## Discussion

Our study found that several environmental factors are influential in the cumulative number of SARS-CoV-2 infection and mortality, with particular factors having apparently significant roles in specific scenarios. Generally, the cumulative number of infections is reduced with increased elevation and solar radiation, but other factors such as poor air quality and population density have significant roles. Surprisingly, NDVI, as a measure of green cover and socioeconomic level in urban settings, was a strong predictive factor of SARS-CoV-2 infection (Table 3).

Previous studies have found that more affluent suburbs in Peru tend to be less populated, have more green space that reflects higher property values (Duarte-Garcia et al., 2018), and therefore, residents may be more likely to have access to private health measures that could decrease rates of infection in the longer term.

Once we separate infection and mortality by ecozones, the influence of environmental factors is similar to looking at Peru as a whole, with a few notable exceptions. Solar radiation is a much more influential factor in the Sierra, where there are strong geographic patterns particularly in the south, and UV radiation is known to be strong in these drier environments (Figure 2). If we take into account the UV radiation as a possible factor that could decrease the survival of the virus in the air, this could account for the decrease of infected people in the sierra and in the south of the country.

Mean NO_2_ concentration is an overriding factor for the cumulative number of mortality for ages below 30. Higher mean NO_2_ concentration is indicative of anthropogenic activity, such as fossil fuel consumption and biomass burning, which occur in more population dense districts, but it is only moderately correlated with population density in Peru. Mean NO_2_ concentration in 2019 was highest in the metropolitan cities of Lima, Arequipa and the small southern city of Moquegua, where NO_2_ density of above 250 µmol m^−2^ were found year round. Seasonal high concentrations, due to agricultural and other biomass burning, are found in cities in the Amazon basin during the dry season. The overall detrimental effects of air pollution on the health and mortality within populations, in accordance to type and length of exposure is known (Carey et al, 2013), along with other studies exploring its effects on the current pandemic (Fattorini and Regoli, 2020).

Unlike male mortality, we found a significant difference in the mean age of women who died across ecozones in Peru, with a higher average age for women in the Sierra. To our knowledge, this is the first study to indicate the multiple factors that seem to influence female mortality, including a negative correlation with elevation and mean solar radiation, but a larger positive correlation with population density, NDVI and mean NO_2_ density. Female mortality from SARS-CoV-2 is lower than males in all the countries possibly due to the expression of the ACE2 (angiotensin□converting enzyme 2) receptor regulated by female hormones. However, protection is lost in older women, as the levels of hormones decrease. The probability of infection in older women becomes similar to their male counterpart, but there is still a higher rate of fatalities in men than women independent of age (Jin Jiam-Min et al, 2020).

Our study also found elevation to be an influential factor in case fatality rate for the coast, sierra, high montane forest, lowland rainforest, and the country as a whole when looking at the data at the district level. This is contrary to the findings Segovvia-Juarez et al. (2020), which studied cases at the provincial level, instead of the district level, and found that the case fatality rate was not modified by elevation. Our analysis indicates a rather weak but statistically significant correlation. However, we acknowledge that further studies still need to be conducted to see if this relationship continues throughout the course of the pandemic.

The limitations of this study are indicative of the asymptomatic nature of SARS-CoV-2 for many patients. It is currently unclear the magnitude of underestimation occurring at the present time and accurate numbers may not become available until widespread molecular testing is performed. Also, news reports indicate that the cumulative number of mortality may also be significantly underestimated due to the lack of testing and patient care in overwhelmed urban and rural hospitals, particularly during the peaks of SARS-CoV-2 infection.

In conclusion, elevation is one of several factors that has determined the number of infections and mortality. Other significant factors include population density, air quality, solar radiation and NDVI, as a measure of both green cover and socioeconomic level. Poor air quality was the single most important factor to determine mortality below the age of 30. We also found that case fatality rate is modified, albeit weakly, by elevation, which is contrary to previously published findings. As more data becomes available, this study can be replicated to see if the relationship between SARS-CoV-2 and climatic and environmental factors are maintained over time.

## Data Availability

All the data is available at https://www.datosabiertos.gob.pe/

## Acknowledgement

The authors wish to thank Professor Dr. Michal Horowitz for her critical reading and discussion of the manuscript.

## ON-LINE DEPOSITORY

### Brian R. Zutta, Diana Flores, Eduardo Rojas, Victor J. Samillan

#### Ecozones

For the purposes of our study, the ecozones originally categories as “selva alta accessible” (accessible high jungle) and “selva alta dificil” (difficult high jungle) were merged to high montane forest ecozone. Similarly, the hydromorphic zone was merged with the lowland rainforest ecozone.

#### Climate data

A set of climate metrics were obtained from WorldClim version 2.1 (Fick & Hijmans, 2017). These metrics are derived from monthly climatologies, which include monthly temperature, rainfall, wind speed, and solar radiation. These monthly climatologies were developed using a series of global and country networks of weather stations and represent a historical baseline from the years 1979 to 2000. The data is interpolated into monthly climate metrics at a 1 km spatial resolution and at the global scale. Eleven temperature and eight precipitation metrics were used, which represent measures of annual means and seasonality. Three wind speed and three solar radiation metrics were also used, which include annual mean, annual standard deviation and annual maximum. The total number of climate layers initially available for this study was 29 before they were reduced to avoid collinearity

#### Environmental data

Environmental data was obtained from a variety of satellite sensors that detect environmental variables, including elevation, vegetation cover, and air quality. We used the Shuttle Radar Topography Mission (SRTM) digital elevation data at the native 90 m spatial resolution.

To quantify vegetation cover, we used the MODIS (Moderate Resolution Imaging Spectroradiometer) Terra 16-day NDVI (Normalized Difference Vegetation Index) product to calculate the mean NDVI value over Peru for the year 2019. NDVI is a widely used spectral index that is highly correlated with vegetation and has a range of value from +1 to –1, where values close to +1, indicate dense green vegetation, and values close to 0 indicate barren ground or artificial surfaces. NDVI values of Peruvian rainforests or well irrigated agricultural plants would approach a value of 1, while cities with mostly artificial surfaces and very few urban trees or gardens would have an average value closer to zero.

We infer long term exposure to air pollution through the metrics of two health-relevant pollutants, which include particulate matter at 2.5 μm (PM 2.5) and nitrogen dioxide (NO_2_). Mean PM 2.5 (dust column mass density in kg m^-2^) was obtained from the MERRA-2 (Modern-Era Retrospective analysis for Research and Applications version 2) using the the Goddard Earth Observing System Model, Version 5 (GEOS-5) with its Atmospheric Data Assimilation System (ADAS), version 5.12.4, which was available through NASA’s Giovanni application (v. 4.34; https://giovanni.gsfc.nasa.gov/). We used the native spatial resolution of 0.5 × 0.625° (approximately 3,850 km^2^) and the annual mean for the year 2019. Tropospheric NO_2_ density (µmol m^−2^) for 2019 was obtained from the Copernicus Sentinel-5P satellite, which holds the TROPOspheric Monitoring Instrument (TROPOMI). We used the native spatial resolution of 7 km and metrics of annual mean and maximum value for the year 2019, which was obtained through Google Earth Engine.

The total number of environmental data layers assessed for this study was 6, which were assessed for collinearity through Superman’s Rho correlation and stepwise linear regression collinearity diagnostics.

